# A novel hyperinflammation clinical risk tool, HI5-NEWS2, predicts mortality following early dexamethasone use in an observational cohort of hospitalised COVID-19 patients

**DOI:** 10.1101/2021.06.16.21259011

**Authors:** Michael R Ardern-Jones, Hang T.T. Phan, Florina Borca, Matt Stammers, James Batchelor, Isabel C. Reading, Sophie V. Fletcher, Trevor Smith, Andrew S Duncombe

**Author notes:** Corresponding author: Michael Ardern-Jones.

## Abstract

**Background:** The success of early dexamethasone therapy for hospitalised COVID-19 cases in treatment of Sars-CoV-2 infection may predominantly reflect its anti-inflammatory action against a hyperinflammation (HI) response. It is likely that there is substantial heterogeneity in HI responses in COVID-19.

**Methods:** Blood CRP, ferritin, neutrophil, lymphocyte and platelet counts were scored to assess HI (HI5) and combined with a validated measure of generalised medical deterioration (NEWS2) before day 2. Our primary outcome was 28 day mortality from early treatment with dexamethasone stratified by HI5-NEWS2 status.

**Findings:** Of 1265 patients, high risk of HI (high HI5-NEWS2) (n=367, 29.0%) conferred a strikingly increased mortality (36.0% vs 7.8%; Age adjusted hazard ratio (aHR) 5.9; 95% CI 3.6-9.8, p<0.001) compared to the low risk group (n= 455, 36.0%). An intermediate risk group (n= 443, 35.0%) also showed significantly higher mortality than the low risk group (17.6% vs 7.8%), aHR 2.2, p=0.005). Early dexamethasone treatment conferred a 50.0% reduction in mortality in the high risk group (36.0% to 18.0%, aHR 0.56, p=0.007). The intermediate risk group showed a trend to reduction in mortality (17.8% to 10.3%, aHR 0.82, p=0.46) which was not observed in the low risk group (7.8% to 9.2%, aHR 1.4, p =0.31).

**Interpretation:** The HI5-NEWS2 measured at COVID-19 diagnosis, strongly predicts mortality at 28 days. Significant reduction in mortality with early dexamethasone treatment was only observed in the high risk group. Therefore, the HI5-NEWS2 score could be utilised to stratify randomised clinical trials to test whether intensified anti-inflammatory therapy would further benefit high risk patients and whether alternative approaches would benefit low risk groups. Considering its recognised morbidity, we suggest that early dexamethasone should not be routinely prescribed for HI5-NEWS2 low risk individuals with COVID-19 and clinicians should cautiously assess the risk benefit of this intervention.

**Funding:** No external funding.

## Introduction

Dexamethasone therapy for COVID-19 is the most significant therapeutic intervention in treatment of severe Sars-CoV-2 infection to date and is supported by clinical trial evidence demonstrating a reduction in mortality as reported by the RECOVERY trial (1) and subsequently confirmed in other studies (2) (3) (4). This is in contrast to the use of glucocorticoids in other severe viral respiratory infections which have a long history, but to date remain controversial and lack evidence from prospective clinical studies. Hyperinflammation (HI), characterised by a rapid increase in systemic release of cytokines such as IL-1 and IL-6, has been reported to explain the association of high fever, high C-reactive protein (CRP), hyperferritinaemia and coagulopathy that are more prevalent in COVID-19 than influenza (5, 6) translating into increased morbidity and mortality. The UK COVID-19 Therapeutics Advice & Support Group (CTAG) on use of immunomodulatory agents in COVID-19 identifies COVID-HI as a specific subgroup of HI syndromes (Supplementary Fig 1) (7, 8). There is consensus that HI syndromes have a better outlook if identified and treated early (9, 10) and the most effective initial intervention is steroid therapy (10, 11).

Whilst current guidance recommends dexamethasone only for severe COVID-19 who are oxygen dependent and hospitalised, it remains unclear whether HI exists in all severe cases of COVID-19 or whether there may be a spectrum of HI within this group. It is possible that responsiveness to dexamethasone may be variable where better responses are seen in patients showing greater degrees of HI ranging to very poor or even adverse responses seen in patients with minimal evidence of HI. Indeed, some well known immediate adverse effects from dexamethasone especially impaired antiviral responses, glucose control, and severe fungal infections(12), have been reported in COVID-19 (13). Indeed, in non-HI cases, other factors are central to mortality such as direct viral invasion of pulmonary tissue (14), existence of significant cardiac (15) and pulmonary (16) co-morbidities and renal failure (17) and require alternative therapeutic strategies. Therefore, targeting the HI group for steroid treatment would seem critically important.

Many algorithms already exist for overall mortality estimation in severe COVID-19 such as ISARIC4 in which the strongest predictor by far is increasing age (18). The National Early Warning Score-2 (NEWS2) (19) has been recommended for assessment of COVID-19 (20). However, no algorithms predict response to steroid therapy.

Therefore, here we set out to assess COVID-19 induced HI, as measured by a novel score (HI5), combined with a validated measure of generalised medical deterioration (NEWS2) to compare treatment response to dexamethasone in HI subgroups. Our primary outcome was 28 day mortality with and without early treatment with dexamethasone stratified by HI5-NEWS2 status.

## Methods

### Ethical considerations

The study followed the principles of the Declaration of Helsinki and was approved by the National Research Ethics Service (Identification of Novel Factors Leading to Activated Macrophage Expansion in COVID19 and related conditions to guide targeted intervention, INFLAME COVID-19 Study, NRES 286016). The study is reported here in accordance with STROBE guidelines (21) and ClinicalTrials.gov ID: NCT04903834.

### Study population

Patients from University Hospitals Southampton NHS Foundation Trust (UHS) were the population for this study. All hospitalised cases of COVID-19 infection that tested positive for SARS-CoV-2 viral RNA in our laboratory between 07/03/2020 and 14/03/2021, n= 2531 were included. We standardised the data with respect to the day of first diagnosis of SARS-CoV2 PCR positivity in our laboratory which we designated as Day 0. Comorbidities were identified from ICD-10 coding extracted from clinical records. The purpose of this study was to develop an early warning system for HI relevant to routine clinical practice. In our cohort rapid access to SARS-CoV-2 PCR testing was available throughout the study, and clinical teams sent samples for PCR as soon as they considered the diagnosis of COVID-19, or routinely on admission. Therefore, to compare patients, we chose to pragmatically normalise all parameters to the date of virus confirmation by SARS-CoV-2 PCR test confirmation.

‘Early’ in the hospitalised disease course was designated Day -1 to 2. 373 individuals >85 years were excluded from the primary analysis (Fig. 1). HI5 and NEWS2 scores were calculated as below based on the most abnormal result over this initial 4 day period (Day-1 to 2). To avoid bias from imputation of missing results, we only included patients with a full dataset of all HI5 parameters (thus excluding a further 427 individuals).

**Figure 1.**
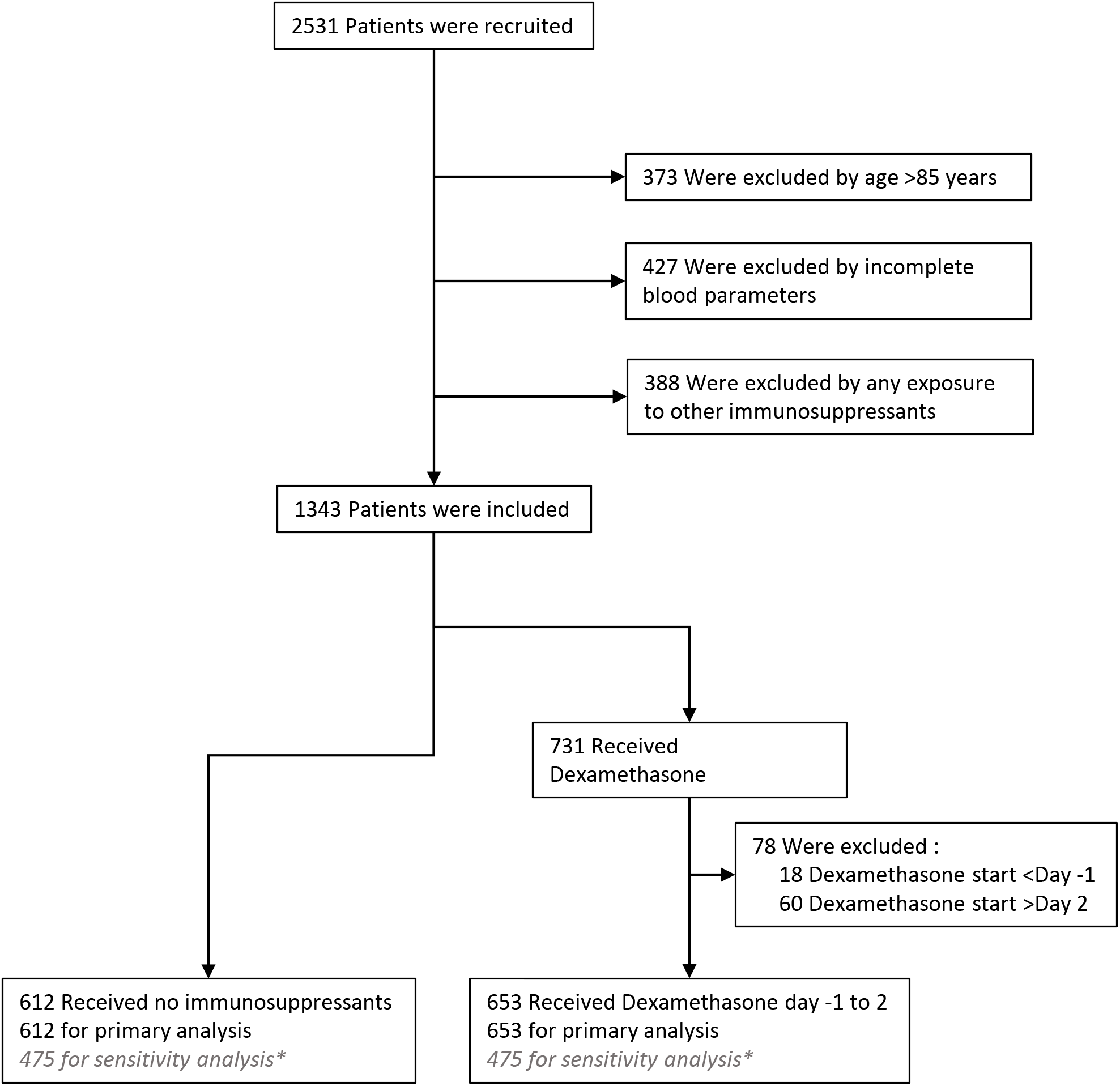
Enrolment, and Inclusion in the Primary Analysis. Electronic records were available for 2531 of 2531 patients (100%) of hospitalised COVID-19 cases. 373 cases were excluded due to age (>85 years), and 427 through incomplete blood parameter measurements. 388 cases received other immunosuppressants as well as or instead of dexamethasone and were excluded. 78 dexamethasone treated cases were excluded because the treatment was started before or after the window of interest.

In response to data released by the UK Chief Medical Officer (16 June 2020) UHS COVID-19 management guidelines were updated immediately to recommend dexamethasone therapy for all hospitalised COVID-19 cases requiring respiratory support with supplemental oxygen to maintain oxygen saturation >94%, Based on data from the Recovery group (1). In order to focus specifically on the intervention of early dexamethasone in these COVID-19 patients, we then excluded patients who were on other immunosuppressive therapy (n=388) and a further group who had received prior dexamethasone at Day<-1 or had received late dexamethasone at >Day2 (n=78, Fig. 1).

Similar to recent COVID-19 studies, the primary outcome utilised in this study was risk of mortality by Day 28 versus survival to Day 28 in patients treated with early dexamethasone (initiated day -1 to 2) compared with those not treated with dexamethasone or any other immunosuppressant.

### Development of predictive score for HI

There is no consensus definition of HI although recognition that COVID-19 induced HI may be considered a subgroup of the overarching term (7, 8). We therefore chose a priori routine laboratory markers of HI known to indicate severity in other clinical contexts. A key driver in our choice of parameters was the common availability of such indicators in routine laboratory practice with rapid result turnaround times to facilitate urgent clinical decision making. HI5 parameter selection excluded parameters necessitating cytokine assays such as IL-6 (22), TNF (23) and GM-CSF (24) that are currently not routinely available in many hospitals. CRP is recognised universally as a key indicator of infection-induced inflammation but confounders such as underlying disease infection make it unreliable as a single indicator (25). Serum ferritin is the most sensitive single indicator for the most severe form of HI, secondary Haemophagocytic Lymphohistiocytosis (sHLH) (26). While complete sHLH criteria are rarely fulfilled in COVID-19 (27, 28) and the degree of elevation in COVID-19 is less than in sHLH, ferritin is still likely to be an important indicator for COVID-HI (29, 30). Unlike most viral infections, COVID-19 induces a neutrophilia (31), a key component of HI in both infective and inflammatory diseases, and therefore this was included as absolute neutrophil count. Absolute lymphocyte count is relevant to HI responses as this may represent virus induced immunosuppression, co-existing disease or concurrent immunosuppressive therapy (7). Platelet count reduction correlates with risk of COVID-19 induced microangiopathic coagulopathy known to associate with HI (32). 5 key parameters were selected: C-reactive protein (CRP), serum ferritin, neutrophil, lymphocyte and platelet absolute counts. While these 5 parameters will not encompass all possible measures of COVID-HI, together they form a coherent and rapidly and universally assessable group of measurements. This novel algorithm was developed from a data cut of UHS COVID-19 cases up to 24-Jun-2020 (n=539) to develop the HI5 algorithm before the widespread use of dexamethasone or other anti-inflammatory agents.

Each parameter was then assessed individually to define thresholds to score 0-4 and weighting added based upon analysis of correlation with the key outcome measure of mortality to produce a total HI5 score out of 44 (presented furthermore as a percentage of maximum score, Table 1). HI5 was made binary to make it easily clinically applicable, and a ‘high’ threshold was set pragmatically by splitting the data in two (approximately 50% of the dataset each) according to the median score and a score of ≥28% selected to classify as high HI5 (n=634) vs low HI5 (n=621).

**Table 1.** The HI5 score algorithm. 5 routinely available blood test parameters are scored based on their value, and weighted. The sum of each parameter score results in the HI5 score. Percentage values are presented.

| HI5 score | 0 | 1 | 2 | 3 | 4 | weight | Total score |
| --- | --- | --- | --- | --- | --- | --- | --- |
| CRP (mg/L) | ≤50 | >50 | >100 | >150 | >250 | 3 | 12 |
| Ferritin (µg/L) | ≤500 | >500 | >1000 | >2000 | >4000 | 2 | 8 |
| ANC (x10<sup>9</sup> /L) | ≤4 | >4 | >8 | >12 | >20 | 3 | 12 |
| ALC (x10<sup>9</sup> /L) | ≥1.5 | <1.5 | <0.9 | <0.6 | <0.3 | 2 | 8 |
| Platelets (x10<sup>9</sup> /L) | ≥250 | <250 | <150 | <100 | <50 | 1 | 4 |
|  |  |  |  |  |  | Max score | 44 |

The National Early Warning Score 2 (NEWS2) for hospitalised patients combines scores for each of 7 routine bedside measurements of physiological parameters to provide an overall NEWS2 score. The following parameters are included: respiratory rate, oxygen saturation, supplemental oxygen, systolic blood pressure, pulse rate, consciousness and temperature. The combination of these values provides a score between 0 and 20 (19). The purpose of the NEWS2 score is to assess acutely ill patients and a score of ≥5 is validated as a threshold to identify deterioration in patients who require intervention (19). Therefore, to identify acutely ill patients, NEWS2 was also examined and a score ≥5 was designated as high risk and NEWS2 <5 designated as low risk.

### Data handling and statistical analysis

Structured and semi-structured data was accrued from the trust integration engine using SQL Developer 4.2 queries and then cleaned/transformed using python 3.7 and associated libraries: *numpy* and *pandas*. Analysis was performed using *matplotlib, seaborn* and *scipy*. Using this approach to retrospectively retrieve data from the electronic hospital record system we collected all blood parameters, bedside observations, and prescribing. In addition, clinical coding information was used to retrieve comorbidity data. Mortality outcome was retrieved from a central NHS Spine database. For the 8/1265 analysed patients where 28 day censoring was not possible (within 28 days of data cut), these cases were censored early with the censor time status, and were indicated on Kaplan-Meier survival curves as indicated. Importantly, patients lacking any parameter of HI scoring system were excluded from the analysis ensuring no bias in HI scoring from imputation of missing data. In UHS, NEWS2 is calculated at the bedside by the clinical team and data input to the electronic patient care record. Complete NEWS2 data was available for 73.8% of cases. However, separately, the dataset contained key data elements of the NEWS2 algorithm including respiratory rate, Oxygen saturations, and temperature (97.1%, 97.1%, and 96.9% of cases respectively). To address the missing NEWS2 data, imputation of NEWS2 from these parameters was undertaken using the K-nearest neighbour method and sensitivity analyses without imputed data reported.

Statistical analysis was undertaken in python 3.7, R (RStudio Version 1.4.1106) and GraphPad, Prism (8.4.3). All data was censored at Day 28, for Kaplan-Meier survival analysis. For the primary outcome of 28-day mortality, the hazard ratio from Cox regression was used to estimate the mortality hazard ratio. To account for any differences in distribution of age across the cohort, all hazard ratios quoted are adjusted using Cox proportional hazards analysis. Unadjusted hazard ratios were estimated using the Log-Rank method. Results without age adjustment are provided. For t-test comparison of demographic measurements between dexamethasone treated and those not treated with dexamethasone, statistical significance was determined using the Holm-Sidak method, with alpha = 0.05. Each characteristic was analysed individually, without assuming a consistent SD. The full anonymised database is held by the research team in the University of Southampton and data linkage is strictly controlled by the data informatics team, University Hospitals Southampton NHS Foundation Trust as per ethical approval.

## Results

A total of 653 patients received dexamethasone between Day -1 and Day 2 (dexamethasone group) and a total of 612 patients did not (untreated group) and there was no statistically significant difference between the groups with respect to age, sex, ethnicity, or comorbidities. Patient age, which is known to be the dominant prognostic factor, showed no overall statistically significant difference between the treated and untreated cohorts (mean age 59.08 versus 61.42 respectively, p=0.08, Table 2). However, to exclude the influence of possible age differences in subgroups, age adjusted analyses are reported throughout (with unadjusted results supplementary). Ethnicity status has been shown to adversely affect outcome in COVID-19 and non-white patients were well represented in both cohorts (26.19% in the dexamethasone treated group versus 25.82% in the untreated group, p=0.99). Key co-morbid drivers of adverse outcome were also not significantly different between the groups. In the treated and untreated groups respectively, chronic lung disease was present in 19.14% versus 21.41%, p=0.90, cardiac co-morbidity was present in 24.04% vs 30.39%, p=0.10, severe renal impairment was seen in 0.77% vs 1.31%, p=0.90, severe liver disease in 1.68% vs 1.80%, p=0.99 and diabetes was present in 16.69% vs 17.81%, p=0.97 (Table 2).

**Table 2.** Characteristics of the Patients with confirmed SARS-CoV-2 severe acute respiratory syndrome coronavirus 2 according to treatment. Plus–minus values are means ±SD. HIV denotes human immunodeficiency virus, NA not applicable, and SARS-CoV-2 severe acute respiratory syndrome coronavirus 2.

| Characteristic |  | Treatment |  | Adjusted p-Value |
| --- | --- | --- | --- | --- |
|  |  | No dexamethasone | Dexamethasone |  |
| Number |  | 612 | 653 |  |
| Age (years) |  |  |  |  |
| | Mean (± SD) | 61.42 ±16.80 | 59.08 ±14.72 | 0.08 |
|  | Distribution |  |  |  |
| Sex - no. (%) |  |  |  | 0.52 |
|  | Male | 345 (56.37) | 398 (60.95) |  |
|  | Female | 267 (43.63) | 255 (39.05) |  |
| Ethnicity - no. (%) |  |  |  | 0.99 |
|  | White | 422 (68.95) | 448 (68.61) |  |
|  | Black, Asian, or minority ethnic group | 158 (25.82) | 171 (26.19) |  |
|  | Unknown | 32 (5.23) | 34 (5.21) |  |
| Previous co-existing disease - no. (%) |  |  |  |  |
|  | Diabetes | 109 (17.81) | 109 (16.69) | 0.97 |
|  | Heart disease | 186 (30.39) | 157 (24.04) | 0.10 |
|  | Chronic lung disease | 131 (21.41) | 125 (19.14) | 0.90 |
|  | Severe liver disease | 11 (1.80) | 11 (1.68) | 0.99 |
|  | Severe kidney impairment | 8 (1.31) | 5 (0.77) | 0.90 |
|  | Tuberculosis | 5 (0.82) | 1 (0.15) | 0.52 |
|  | HIV infection | 2 (0.33) | 3 (0.46) | 0.98 |

As expected, the group treated with dexamethasone predominantly comprised the second wave of COVID-19 infection (Supplementary Fig 2). Dexamethasone 6mg daily by mouth or intravenously, for 10 days or until discharge was prescribed in accordance with recommendations from the Recovery trial (1).

Mortality at 28 days in hospitalised COVID-19 patients not treated with dexamethasone (started day -1 to 2) was significantly higher in cases with HI5 high risk score measured early in the disease course with deaths in 71 out of 264 patients (26.9%) compared to 35 out of 348 (10.1%) with low HI5 scores (age adjusted hazard ratio (aHR) 2.7, 95% Confidence interval (CI) 1.80-4.10, p<0.001, Fig 2a). For High NEWS2 (score ≥ 5) mortality in dexamethasone untreated cases was also significantly higher with deaths in 63 out of 205 patients (31.7%) versus 43 out of 407 (10.6%) in those with low NEWS2 scores (aHR 3.7, CI 2.5 – 5.50, p<0.001, Fig 2b). NEWS2 and HI5 were developed to predict acute risk of medical deterioration and HI respectively. Although some overlap may exist in some patients, to examine their interrelationship, linear regression analysis of NEWS2 and HI5 showed that their correlation was low (r^2^ = 0.171, Supplementary Fig 3), suggesting that high HI5 or NEWS2 scores independently confer an excess mortality risk over low scores. Therefore, we postulated that combining the two scores may offer a superior predictive tool as compared to either alone. Indeed, high risk individuals (with both high HI5 and high NEWS2 scores) showed a greater mortality 36.0% (50/139) than observed 7.8% (22/282) in low risk cases (low HI5 and low NEWS2) (aHR 5.9, 95% CI 3.66-9.8, p <0.001, Fig 2c,d). The groups with high HI5 or high NEWS2 (not both), showed intermediate mortality (16.8%, 21/125; and 19.7%, 13/66 respectively) and Cox regression analysis showed no statistical difference between these two groups (p=0.64). These two groups were therefore subsequently classified as intermediate risk. As compared to low risk groups, intermediate risk groups showed a higher mortality (aHR 2.2, CI 1.3-3.7, p=0.005, Fig 2d). Sensitivity analyses without adjustment for age, and for the effect of NEWS2 imputation, resulted in similar findings (Supplementary Table 1 and Supplementary Figure 4 respectively).

**Fig 2.**
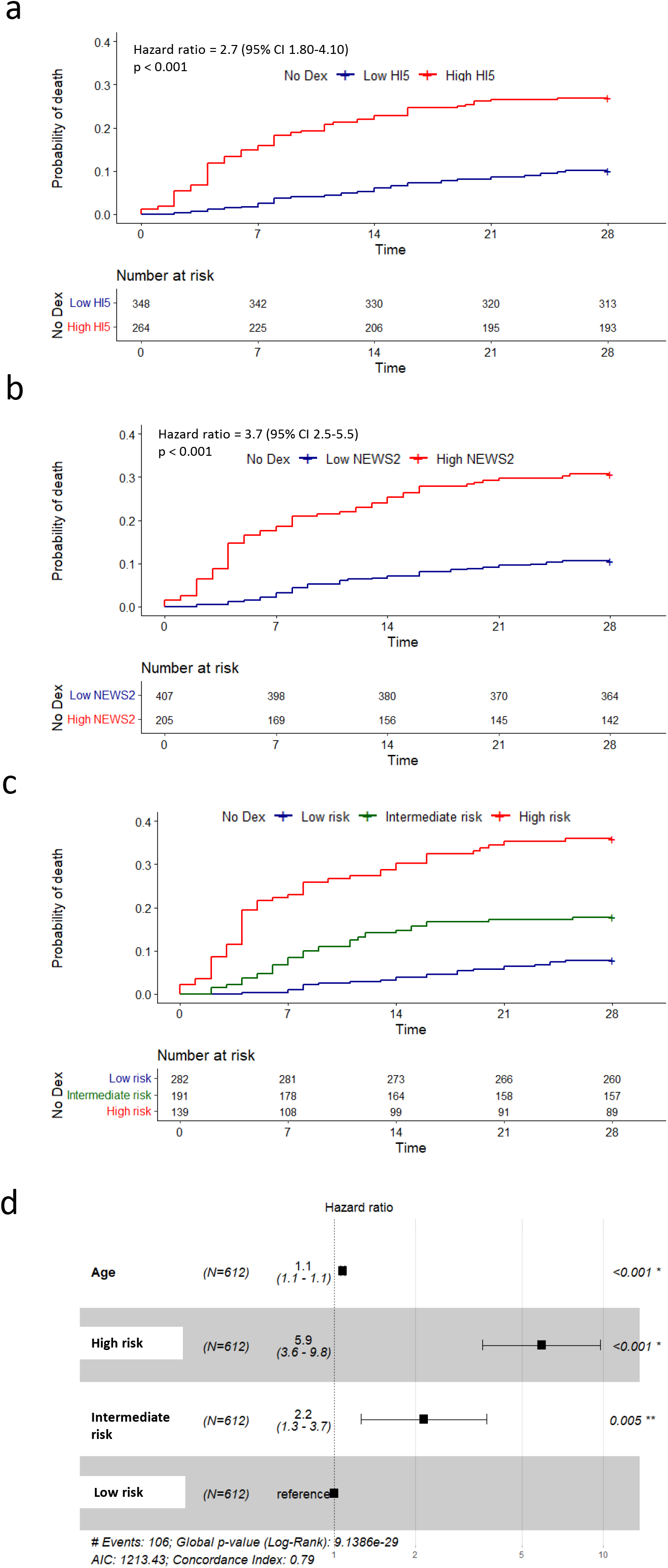
Mortality at day 28 in cases not treated with dexamethasone. Kaplan–Meier survival curves for 28-day mortality among patients who were not treated with dexamethasone with high (red line) or low (blue line) HI5 scores (a) or NEWS2 scores (b). Panel C, Kaplan-Meier survival curves for cases classified as high risk (High HI5 and High NEWS2,, red line), intermediate risk (High HI5 or High NEWS2,, green line), and low risk (Low HI5 and Low NEWS2,, blue line). (d) Cox regression analysis for Age (per year of life), and HI5-NEWS2 risk status. All quoted hazard ratios are adjusted for age (aHR). At risk data are listed beneath plots. Time measured in days.

To examine the effect of dexamethasone in COVID-19 in a real-world population we compared survival in the early dexamethasone treated versus untreated patients. Treatment of COVID-19 with early dexamethasone conferred a modest but non-significant reduction in mortality (12.7% vs 17.3%, aHR 0.93, CI 0.7-1.2, p=0.62, Fig 3a) in our entire cohort. Since a major component of the action of dexamethsaone is anti-inflammatory, and is likely to reduce HI in COVID-19, we examined whether the benefit from dexamethasone was stratified by low, intermediate or high HI5-NEWS2 risk status as measured at day -1 to 2. Strikingly, in the HI5-NEWS2 high risk group, treatment with dexamethasone significantly reduced day 28 mortality from 36.0% to 18.0% (aHR 0.56, CI 0.37-0.85, p=0.007, Fig 3b). In the intermediate risk group, a non-significant reduction in mortality was observed: 17.8% to 10.3% (aHR 0.82, CI 0.49 to 1.4, p=0.46, Fig 3c). In the low risk group, treatment with dexamethasone associated with a non-significant increase in mortality (7.8% to 9.2%) (aHR 1.4, CI 0.73 to 2.6, p=0.32, Fig 3d). Sensitivity analyses without imputation for missing NEWS2 data, resulted in similar findings (Supplementary Figure 5).

**Fig 3.**
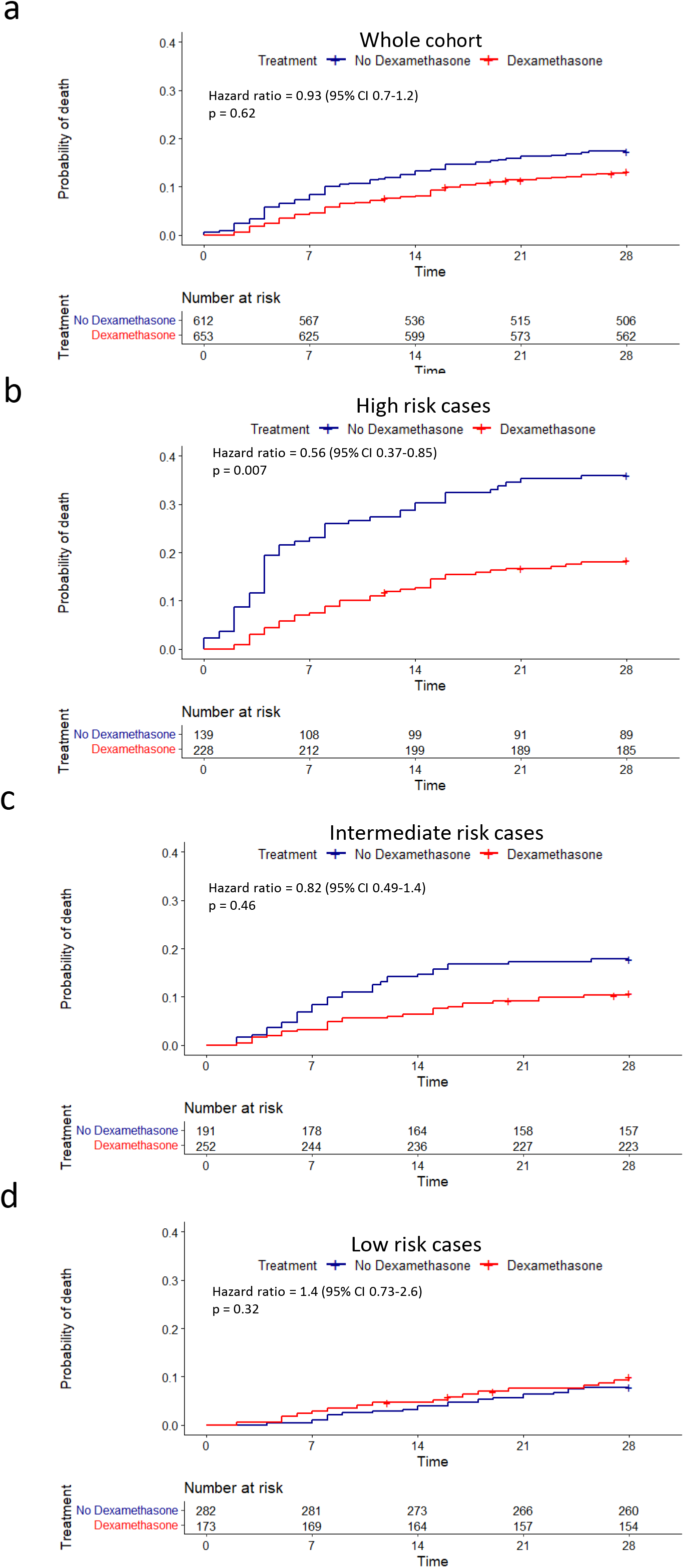
Effect of dexamethasone on mortality at day 28. (a) Kaplan–Meier survival curves for 28-day mortality among the whole cohort in those who were treated with dexamethasone (red line) vs untreated cases (blue line). (b-d) Kaplan–Meier survival curves for those treated with dexamethasone (red lines) vs untreated (blue lines) in HI5-NEWS2 high risk (b), intermediate risk (c) or low risk (d) groups. All quoted hazard ratios are adjusted for age. At risk data are listed beneath plots. Time measured in days. Cases censored before 28 days indicated by +.

## Discussion

We demonstrate that it is possible to classify COVID-19 patients on admission into high, intermediate, and low risk groups for HI, and that HI risk status predicts the benefit from early dexamethasone therapy on mortality. The use of our combined HI5-NEWS2 algorithm demonstrates that patients who have evidence of HI and are acutely ill have the best response to dexamethasone whereas those who lack evidence of HI and are relatively well have no response and may risk steroid related morbidity.

Whilst there are published tools for COVID-19 outcome prediction (18), multiparameter models do not distinguish between the pathophysiological pathways leading to the adverse outcome. In addition, advancing age is such a dominant predictor of survival in COVID-19 (18), that important subgroups may be missed if mortality is used as a primary outcome measure in a non-hypothesis driven approach. Therefore, we used a pre-defined measure specifically designed to measure hyperinflammation and adjusted for the influence of age throughout. The attractiveness of identifying patients showing HI features is the relevance to therapies targeting HI which provides the opportunity to validate a proposed HI algorithm as we have shown. While dexamethasone may have other therapeutic actions in COVID-19 such as ACE receptor targeting, its efficacy as initial therapy in other HI syndromes not involving a viral aetiology suggests that its anti-inflammatory action is a crucial component of efficacy. A recent systematic review reported measurement of hyperinflammation but did not validate for early risk detection or against an anti-inflammatory intervention as we show here (33). We believe early assessment following admission is crucial for rapid clinical decision making so our score was developed for assessment within 2 days of virus confirmation.

The nature of the rapid reporting during COVID-19 imbues various limitations which are relevant to retrospective analyses such as ours. However, whilst potential pitfalls of bias exist, we have minimised the most important forms of bias as follows: assessment was made of 100% of sequential patients admitted to our institution and the management of COVID-19 remained stable throughout the pandemic. By sampling a single institution, clinical teams and the facilities at the institution were the same for both groups. Furthermore, no differences between dexamethasone and untreated groups were identified including all key potential confounding parameters of ethnicity, sex, diabetes, heart disease, and respiratory disease. Indeed, in our whole cohort analysis the benefit from dexamethasone was small, suggesting that any bias in dexamethasone selection would be unlikely to explain the large subgroup differences associated with high or low risk HI5-NEWS2 status.

The original randomised clinical trial from the RECOVERY Collaborative Group (1) showed mortality across all ages for those who received usual care of 25.7% versus 22.9% in those treated with dexamethasone. Our cohort excluded patients ≥85 years and so showed lower overall mortality: usual care was 18.4% (number at risk 612) versus 16.3% in dexamethasone treated groups (number at risk 653) although the trend was similar. Importantly, our results suggest that the benefit first identified by the Recovery trial for dexamethasone treatment in COVID-19 is principally restricted to individuals with HI and not the whole cohort. We show that a simple algorithm that can be used rapidly at the bedside in routine clinical practice can identify early in the admission which patients are most likely to benefit from intervention with early dexamethasone. Furthermore, although our data demonstrate that with dexamethasone treatment of HI5-NEWS2 high risk groups mortality is reduced from 36.0% to 18.0%, this is still twice as high as those with low risk scores. This finding raises the important question of whether targeting the high risk HI5-NEWS2 group with more intensive anti-inflammatory therapies such as with tocilizumab or other early interventional immunosuppressive treatments could reduce their mortality still further. We suggest that a randomised clinical trial of intensified immunosuppression specifically in this HI5-NEWS2 high risk subgroup is warranted to further improve outcomes.

We did not set out to look at morbidity from dexamethasone. Nevertheless, it is concerning that no survival advantage was identified from early dexamethasone treatment in HI5-NEWS2 low risk and intermediate risk groups of hospitalised COVID-19 patients who make up 68.8% of our cases. Indeed, in the low risk group (36% of total) the trend is towards harm in the dexamethasone treated group (Fig. 3d). This could be due to the action of dexamethasone reducing host immune inhibition of viral replication overcoming any benefit from an anti-inflammatory effect in individuals with little evidence of inflammation. This concern has certainly been a factor in the poor overall results of steroid therapy in other viral conditions unless effective anti-viral therapy is given in conjunction. In addition, there is a valid concern supported by anecdotal experience that these individuals may suffer the well-established adverse effects of high dose steroid therapy, including a significant increased risk of new-onset diabetic ketoacidosis (34). New therapies are required in this HI5-NEWS2 low risk group to improve outcomes.

While awaiting these risk stratified trials, we urge caution in prescribing early dexamethasone therapy in COVID-19 in the HI5-NEWS2 low risk group and encourage careful consideration of the potential for harm with respect to steroid induced morbidity.

## Data Availability

Data will be disclosed only upon request and approval of the proposed use of the data by the chief investigator of the INFLAME study, and in alignment with the relevant governance and ethical approval.

## Declaration of interests

No financial support or other benefits from commercial sources for the work reported on in the manuscript, or any other financial interests are declared. No potential conflict of interest or the appearance of a conflict of interest with regard to the work declared.

## Conflict of interest statements

None of the authors declare any conflicts of interest relevant to this work.

## Funding source

No external funding received.

## Acknowledgments

We would like to acknowledge Prof S. Holgate, University of Southampton, for his critical review and advice on manuscript preparation, as well as Dr Anastasia Koutalopoulou and Dr Ying Teo for clinical support.

## Supplementary Figures

**Supplementary Figure 1.**
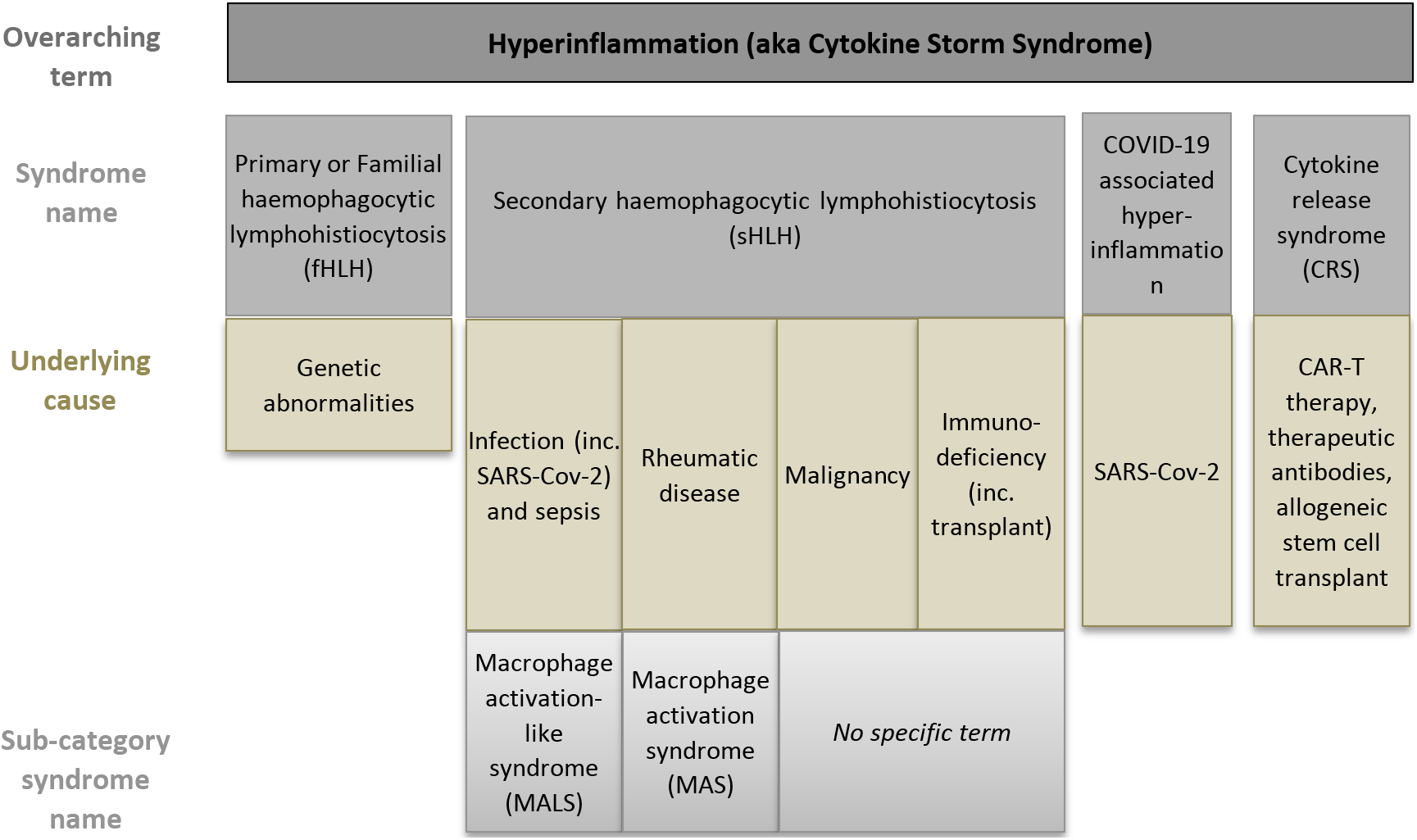
Overview of hyperinflammatory syndromes and their classification (35).

**Supplementary Table 1.** Unadjusted hazard rates (log-rank method) for all cases, High HI5, High NEWS2, and HI5-NEWS2 High risk, Intermediate risk and Low risk cases, versus age adjusted analysis (Cox regression).

| Subgroup | No. of patients (%) |  | Hazard ratio (95% CI) |  |
| --- | --- | --- | --- | --- |
|  | Dexamethasone<br>(N=653) | Usual care<br>(N=612) | Age-adjusted<br>(Cox regression) | Unadjusted (Log-Rank method) |
| All cases | 653 (100.00) | 612 (100.00) | 0.93 (0.7-1.2) | 0.73 (0.55-0.97) |
| High risk HI5-NEWS2 | 228 (34.92) | 139 (22.71) | 0.56 (0.37-0.85) | 0.44 (0.29-0.66) |
| Intermediate risk HI5-NEWS2 | 252 (38.59) | 191 (31.21) | 0.82 (0.49-1.4) | 0.57 (0.34-0.95) |
| Low risk HI5-NEWS2 | 173 (26.49) | 282 (46.08) | 1.4 (0.73-2.6) | 1.3 (0.68-2.4) |

**Supplementary Figure 2.**
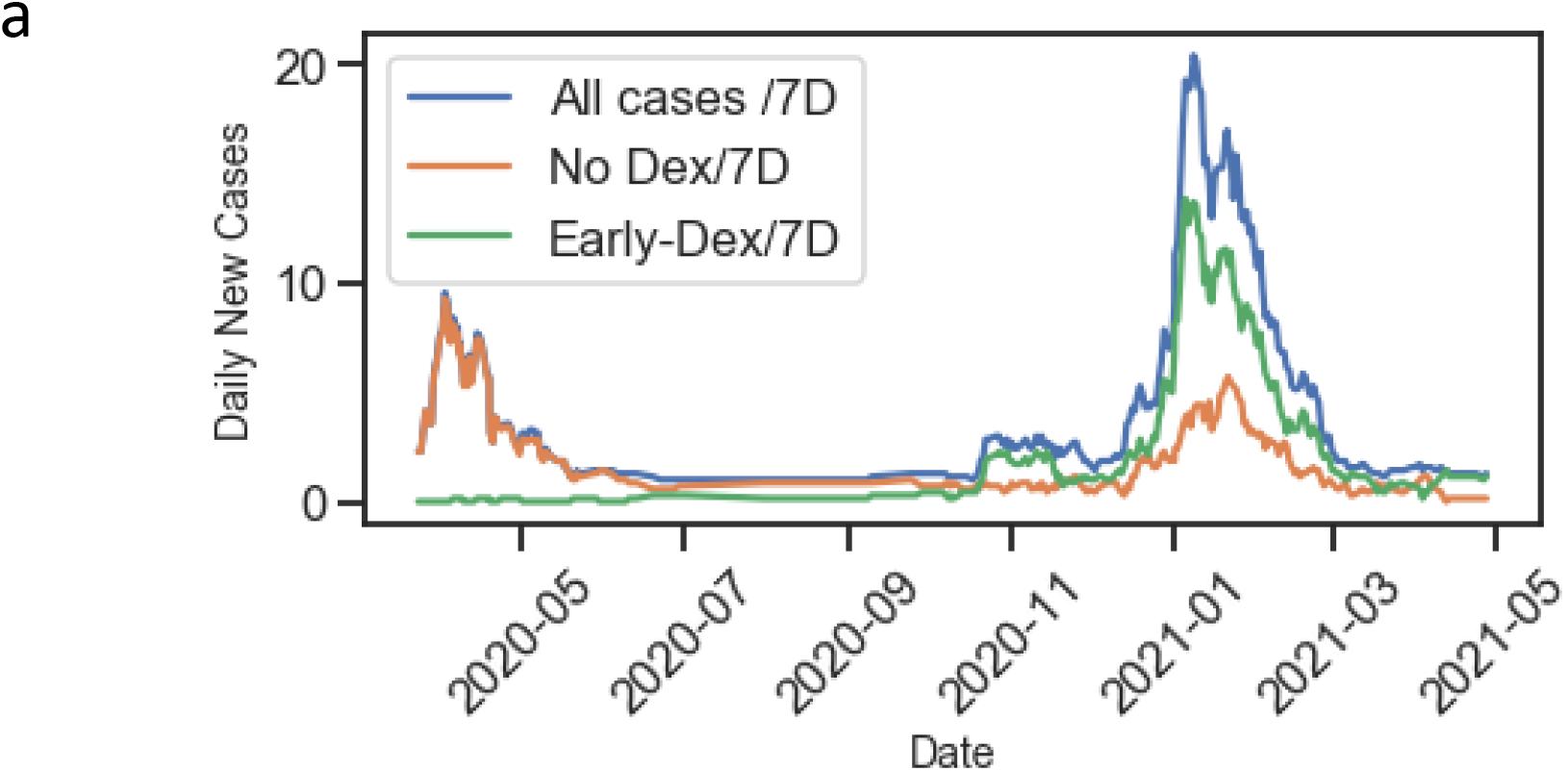
The admission of COVID-19 cases by laboratory confirmed (Sars-CoV-2) polymerase chain reaction confirmation date (X-axis), vs 7 day rolling average of number of cases (Y-axis). Blue line, all cases admitted to the institution; Orange line, cases not treated with dexamethasone recruited in this study; green line, cases receiving prescription for dexamethasone within day -1 to 2 of virus confirmation.

**Supplementary Figure 3.**
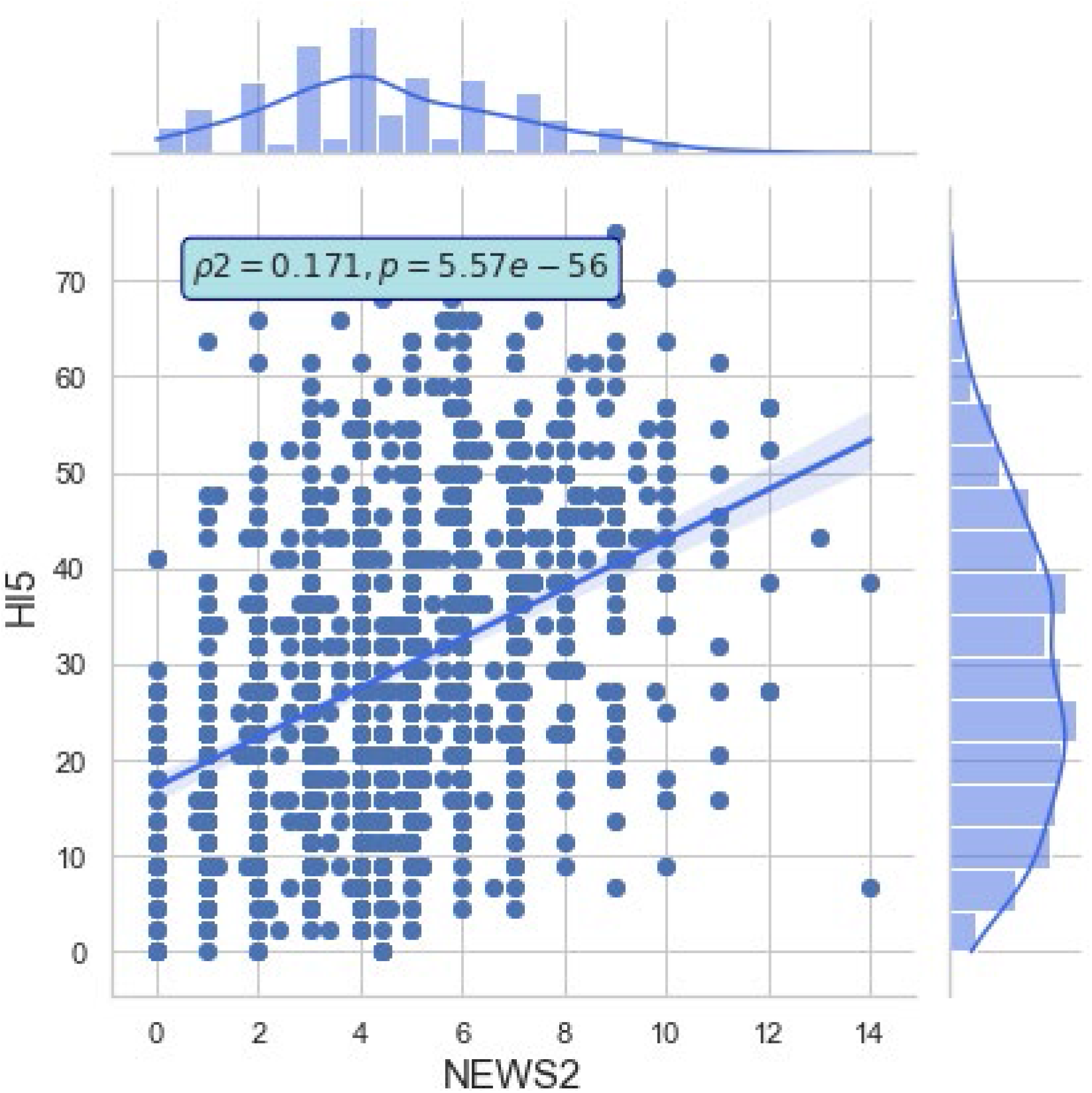
Correlation between HI5 (x-axis) and NEWS2 (y-axis). Pearson’s correlation ρ^2^ reported in the figure.

**Supplementary Figure 4.**
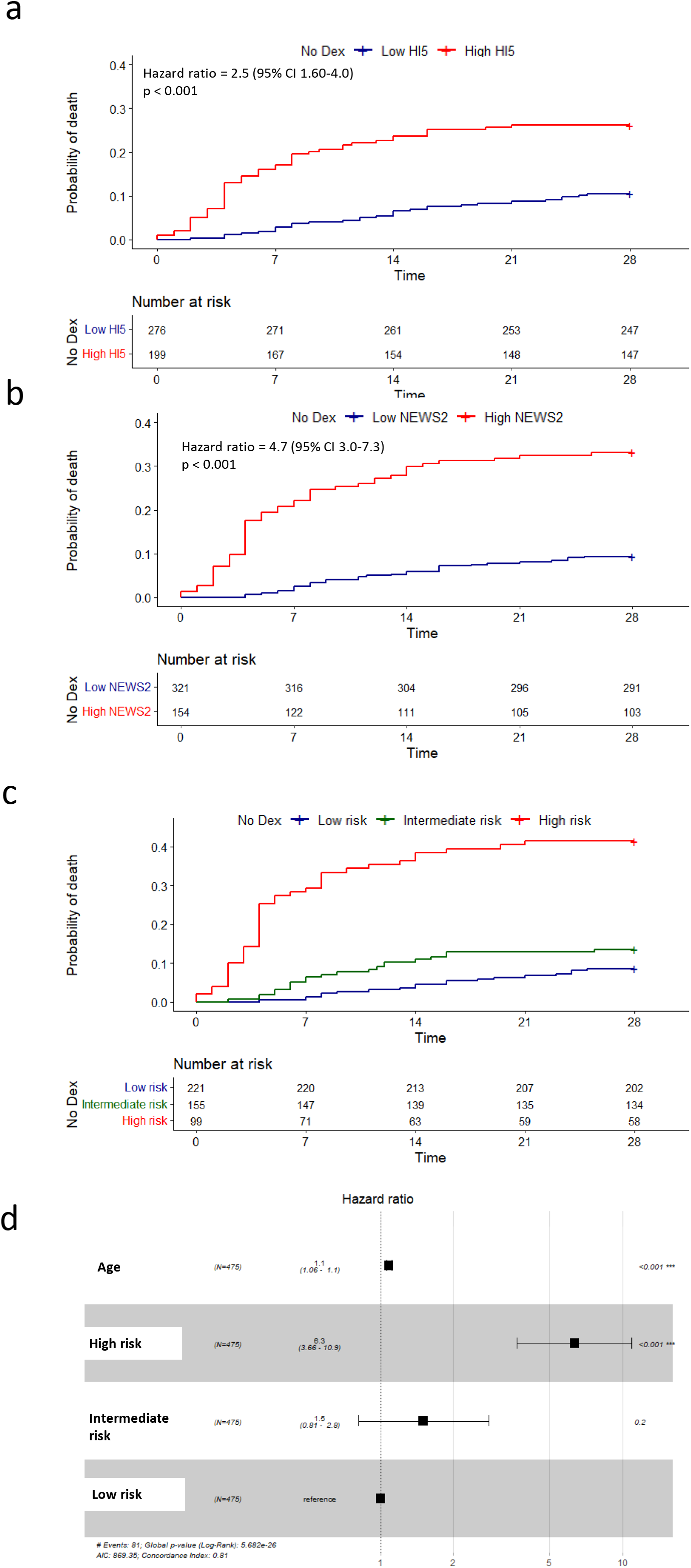
Sensitivity analysis with non-imputed NEWS2 data showing mortality at day 28 in cases not treated with dexamethasone. Kaplan–Meier survival curves for 28-day mortality among patients who were not treated with dexamethasone with high (red line) or low (blue line) HI5 scores (a) or NEWS2 scores (b). Panel C, Kaplan-Meier survival curves for cases classified as high risk (High HI5 and High NEWS2,, red line), intermediate risk (High HI5 or High NEWS2,, green line), and low risk (Low HI5 and Low NEWS2,, blue line). (d) Cox regression analysis for Age, and HI5-NEWS2 risk status. All quoted hazard ratios are adjusted for age. At risk data are listed beneath plots. Time measured in days. Cases censored before 28 days indicated by +.

**Supplementary Figure 5.**
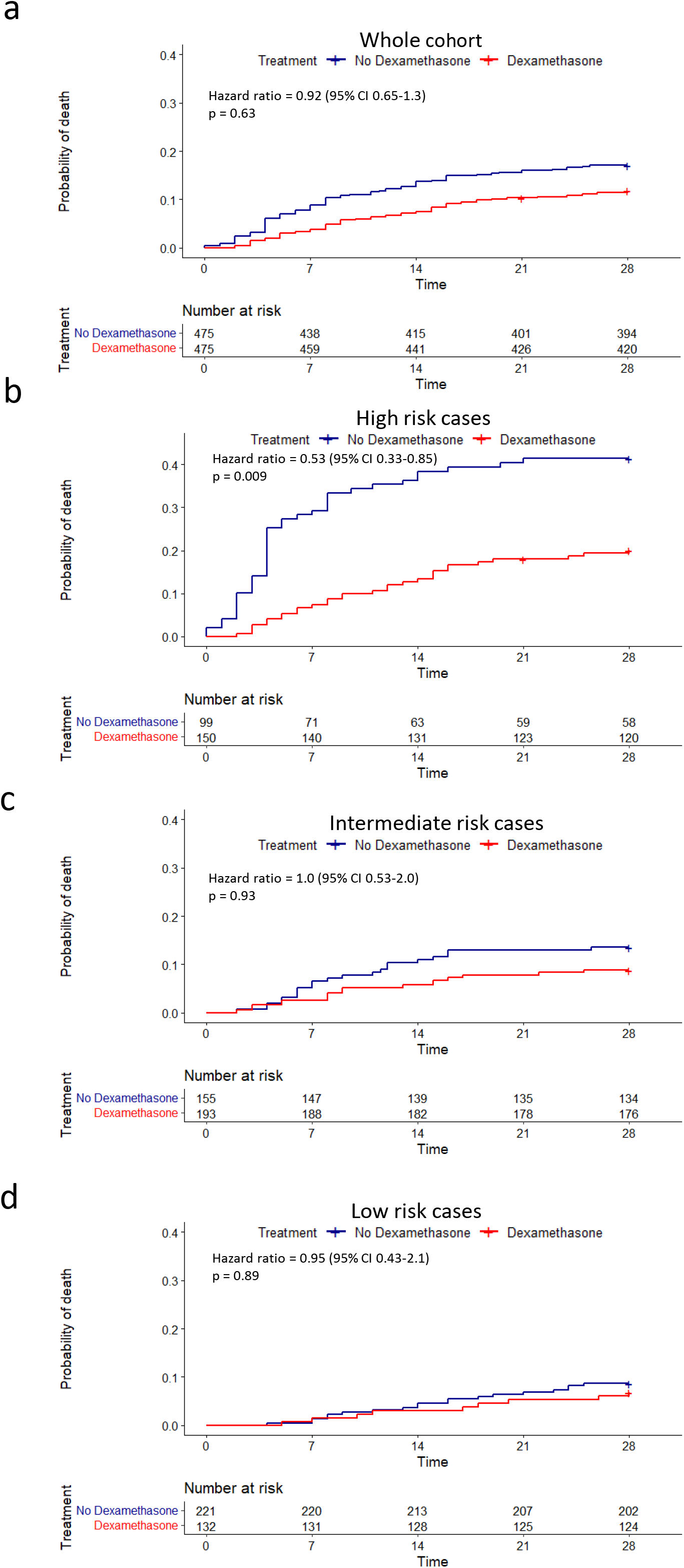
Sensitivity analysis with non-imputed NEWS2 data showing effect of dexamethasone on mortality at day 28. (a) Kaplan–Meier survival curves for 28-day mortality among the whole cohort in those who were treated with dexamethasone (red line) vs untreated cases (blue line). (b-d) Kaplan–Meier survival curves for those treated with dexamethasone (red lines) vs untreated (blue lines) in high risk (b), intermediate risk (c) or low risk (d) groups. All quoted hazard ratios are adjusted for age. At risk data are listed beneath plots. Time measured in days.

## Author contributions

AD and MAJ designed the research study

MAJ, MS, HP, FB, performed the research

HP, FB, JB, MAJ contributed essential informatics tools

MAJ, MS, HP, AD, IR analysed the data

MAJ wrote the first draft of the paper

MAJ, MS, HP, FB, SF, IR, JB, TS, AD contributed to the final manuscript

## Conflict of interest statements

None of the authors declare any conflicts of interest relevant to this work.

## Funding source

No external funding received.

## Ethical approval

UK NHS ethical approval from Health and Care Research Wales, IRAS 286016

https://www.hra.nhs.uk/planning-and-improving-research/application-summaries/research-summaries/inflame-covid19-study-covid-19/

Clinicaltraisl.gov ID: NCT04903834

## Notes

### Competing Interest Statement

The authors have declared no competing interest.

### Clinical Trial

NCT04903834

### Funding Statement

No funding

### Author Declarations

The study followed the principles of the Declaration of Helsinki and was approved by the National Research Ethics Service (Identification of Novel Factors Leading to Activated Macrophage Expansion in COVID19 and related conditions to guide targeted intervention, INFLAME COVID-19 Study, NRES 286016).

